# Immunogenicity and safety of the CoronaVac inactivated SARS-CoV-2 vaccine in people with underlying medical conditions: a retrospective study

**DOI:** 10.1101/2022.04.28.22274402

**Authors:** Chunmei Li, Ao Li, Hanfang Bi, Jun Hu, Fan Yang, Taicheng Zhou, Yupeng Liang, Wei Su, Tianpei Shi, Mei Yang, Rong Wang, Wanting Qin, Zumi Zhou, Jia Wei, Zhenwang Fu, Zijie Zhang

## Abstract

**Background:** People living with chronic disease, particularly seniors older than 60 years old, are lagging behind in the national vaccination campaign in China due to uncertainty of safety and effectiveness. However, this special population made up of most severe symptom and death cases among infected patients and should be prioritized in vaccination program. In this retrospective study, we assessed the safety and immunogenicity of the CoronaVac inactivated vaccines in people with underlying medical conditions to address the vaccine hesitation in this special population.

**Methods:** In this cohort study, volunteers aged 40 years and older, had received two doses of CoronaVac inactivated vaccines (3-5 weeks interval), been healthy or with at least one of the six diseases: coronary heart disease (CAD), hypertension, diabetes mellitus (DM), chronic respiratory disease (CRD), obesity and cancer, were recruited from 4 study sites in China. The primary safety outcome was the incidence of adverse events within 14 days after each dose of vaccination. The primary immunogenic outcome was geometric mean titer (GMT) of neutralizing antibodies to living SARS-CoV-2 virus at 14-28 days, 3 months, and 6 months after full two-dose vaccination. This study is registered with ChiCTR.org.cn (ChiCTR2200058281) and is active but no longer recruiting.

**Findings:** Among 1,302 volunteers screened between Jul 5 and Dec 30, 2021, 969 were eligible and enrolled in our cohort, including 740 living with underlying medical conditions and 229 as healthy control. All of them formed the safety cohort. The overall incidence of adverse reactions was 150 (20.27%) of 740 in the comorbidities group versus 32 (13.97%) of 229 in the healthy group, with significant difference (P=0.0334). The difference was mainly contributed by fatigue and injection-site pain in some groups. Most adverse reactions were mild (Grade 1). We did not observe any serious adverse events related to vaccination. By day 14-28 post vaccination, the seroconversion rates and GMT of neutralizing antibody showed no significant difference between disease group and healthy group, except CAD group (P=0.03) and CRD group (P=0.04) showed slight reduction. By day 90, the neutralizing antibody GMTs were significantly reduced in each group, with no significant difference between diseases and healthy group. By day 180, the neutralizing antibody continued to decrease in each group, but with slower declination.

**Interpretation:** For people living with chronic disease especially seniors older than 60 years, the CoronaVac vaccines are as safe as in healthy people. Although the immunogenicity is slightly different in subgroup of some diseases compared with that of the healthy population, the overall trend was consistent. Our findings highlight the evidence to address vaccine hesitancy for seniors and people living with chronic diseases.

**Funding:** Yunnan Provincial Science and Technology Department (202102AA100051 and 202003AC100010, China), Sinovac Biotech Ltd (PRO-nCOV-4004).

## Introduction

The coronavirus disease 2019 (COVID-19) caused by severe acute respiratory syndrome coronavirus 2 (SARS-CoV-2) continues to result in large number of deaths and remain to be the greatest challenge for public health. As of March 25, 2022, 475 million COVID-19 cases and more than 6 million deaths were confirmed worldwide^1^. Vaccine is considered as a safe and effective way to reduce SARS-CoV-2 infection, COVID-19 mortality and morbidity. Yet, recently emerged Omicron (B.1.1.529 & BA.2), which exhibited strong immune escaping from immunity elicited by natural infection or vaccination, has been rapidly spreading and replaced Delta as the predominant variant worldwide.

Omicron variant has greater transmissibility but causes less severe disease than other latest variant of concern (VOC). Preliminary data suggest that Omicron causes milder symptom and most patients recovered without specific treatment, particularly for young people^2-4^. This is largely thanks to the protection against severe symptom by vaccinations^5,6^. Nevertheless, if infections surge fast in areas with dense population (e.g., metropolitan areas in China), the number of mortality and morbidity would still be non-negligible given the size of population. Moreover, there are over half of seniors aged 60 years or older remain unvaccinated as of March 2022, which may further increase the mortality cases at outbreaks^7^. Death cases analysis in recent wave of outbreak in Hongkong showed that most fetal cases were senior unvaccinated person with chronic diseases^8^. In many countries, senior people with medical conditions are prioritized for vaccination due to extra vulnerability^9-14^. In China, the vaccination among older people with underlying medical conditions were lagging behind due to uncertainty of safety and effectiveness^15-17^. Significant proportion of this vulnerable population had not yet received one or two doses of COVID-19 vaccine. Lessons from the recent wave of Omicron pandemic indicate increasing coverage of vaccination among seniors ≧60 years of age and people with underlying medical conditions are the key to reduce COVID-19-associated morbidity and mortality, which in turn contribute to the global epidemic prevention and control.

There have been lots of studies showing comparable safety and effectiveness of SARS-CoV-2 vaccine in special population (e.g., diabetes, solid organ transplant, autoimmune disease patients), which greatly promoted the vaccination in senior people with and without underlying diseases^18-21^. Systematic evaluation of inactivated SARS-CoV-2 vaccine safety and effectiveness in people with common diseases has been rare. To address vaccine hesitation in seniors and with underlying medical conditions^22,23^, we performed a retrospective study to profile the immunogenicity and safety of CoronaVac (one of the most widely administered inactivated SARS-CoV-2 vaccines in China and many countries) in people with 6 types of chronic diseases.

## Methods

### Study design and participants

We conducted a multicenter retrospective clinical trial in four different study sites (Haikou city, Wenchang city, and Qionghai city, Hainan Province; Kunming city, Yunnan Province; China), aiming to evaluate the immunogenicity and safety of CoronaVac vacancies in people with underlying medical conditions in comparison with age matched healthy individuals.

We recruited participants who have received 2 doses of CoronaVac inactivated vaccine with 3-5 weeks of dose interval and were at the 14th-28th day after the second dose at the time of enrollment. Participants were eligible if they were 40 years of age or older, healthy or diagnosed with any of the 6 most common chronic diseases (hypertension, diabetes mellitus (DM), acute coronary disease (ACD), chronic lung disease (CRD), obesity, and cancer), and were able to understand and complete questionnaires. They will be excluded according to established criteria: had been infected by SARS-CoV-2; had received non-CoronaVac vaccine; with severe mental and neurological diseases; with any other factors unsuitable for clinical observation.

Sex matched healthy participants were recruited as the control group. Since underlying disease condition is common in older adults (≥60 years), participants were grouped into adult (40-59 years old) and senior (≥60 years) subgroups to evaluate the effect of age more accurately on the immunogenicity and safety of the inactivated SARS-CoV-2 vaccine. The disease group was further divided into 6 subgroups based on the 6 common diseases we concerned.

Written informed consent was obtained from each participant before enrolment. The clinical trial protocol and informed consent form were approved by the Committee on Human Subject Research and Ethics of Yunnan University (CHSRE2021021). This study was conducted in accordance with the requirements of Good Clinical Practice of China and the International Conference on Harmonization.

### Procedures

The basic disease information of participants, including disease name, duration, severity and control status, was collected through questionnaires. Participants who came for serum tests 14-28 days after full vaccination were assessed for adverse reaction events within vaccination period. Participant were first screened for grade 3 (daily activity significantly affected, medical attention required, and hospitalization may be necessary) or grade 4 (potential life threat, daily activity severely limited and hospitalization required) adverse reaction events by researcher. Participants were then requested to fill a survey to report grade 1 (short or mild reactions without interfering daily activity) and grade 2 (daily activity mildly interfered, simple treatment or no treatment was necessary). The reported adverse events were graded according to the China National Medical Products Administration guidelines^24^.

Venous blood was collected for neutralizing antibody response assay at day 14-28, 90 and 180 post full vaccination. For all participants, serum samples were collected by Vacutainer SST tubes (BD, USA). For participants at two sites (Haikou and Kunming), we preserved whole blood using Vacutainer EDTA tubes (BD, USA) at room temperature for no longer than 6 hours before isolation. Then we separated PBMCs by density-gradient sedimentation using Ficoll (GE) reagent and cryopreserved at -80°C until testing.

The neutralizing antibodies to live SARS-CoV-2 were quantified using the gold standard of antibody titration: microcytopathogenic effect assay with live virus to quantify the neutralizing antibody in 95 third dose recipients (subjects of live virus test were selected according to the availability of serum sample). 50 μl serum was first inactivated at 56°C for 30 minutes and serially diluted with cell culture medium in two-fold steps. The diluted serum was then incubated with equal volume of live SARS-CoV-2, including wild type (virus titers: 6.0 lgCCID50/mL; passage: P5; GenBank number: MT407649.1) and Delta strains (virus titers: 6.2 lgCCID50/mL; passage: P2; GISAID number: Delta EPI_ISL_1911197), for 2 hours at 36.5°C. Vero cells were then added to the serum-virus mix and incubated at 36.5°C for 5 days. Neutralizing antibody titer was calculated by the dilution number of 50% protective condition^25^.

### Outcomes

The primary safety endpoint was the occurrence of adverse reactions within 14 days after the first dose and the second dose of the vaccination. The primary immunogenic endpoints were the seropositive rate and the geometric mean titers (GMTs) of neutralizing antibodies to live SARS-CoV-2 virus (wild type) 14-28 days, 90 days and 180 days after full two-dose vaccination.

### Statistical analysis

We assessed the safety endpoints in the safety population and assessed the immunogenic endpoints in the participants who completed blood collection at day 14-28, 90 and 180 post vaccination, respectively, and has successful antibody measurements.

For participants enrolled and completed the assays in each group, the age, weight, BMI were described by mean and standard deviation; the gender and nationality were described by the ratio among total participants. For immunogenicity evaluation, we used descriptive statistics (geometrical mean and 95% confidence interval) to summarize antibody levels, and the GMT of post-immunization neutralizing antibody was analyzed after logarithmic conversion, and the least square mean of GMT of post-immunization neutralizing antibody and the ratio between groups and its 95% confidence interval (95%CI) were calculated for comparison. The positive rate of neutralizing antibody after immunization was calculated for the experimental group and the control group, the bilateral 95%CI was calculated by Clopper-Pearson method, and the difference between groups was statistically tested by Chi-square test /Fisher exact probability method. At the same time, geometric mean and 95%CI were used to statistically describe the GMT of the immune neutralizing antibody of participants in each cohort, and logarithmic converted group T test was used to statistically test the difference between groups.

For safety evaluation, in accordance with the protocol, systemic adverse events and local adverse events will be classified and counted. In this study, Treatment Emergent Adverse Events (TEAE) occurred after inoculation were statistically analyzed. Adverse events were expressed by counts and frequency. The number and incidence of all adverse events in each group were calculated, and the differences between groups were statistically compared using Fisher’s exact test method. Descriptive statistics were made for the severity, dosage, and occurrence time of adverse events. The adverse events after each dose of inoculation were statistically analyzed. Adverse events for each dose were analyzed based on the safety population of each dose. No serious adverse event has been reported in this study and thus has not been analyzed.

All statistical analyses were performed by R scripts. This trial is registered with ChiCTR.org.cn, ChiCTR2200058281.

## Results

Between Jul 5 and Dec 30, 2021, we recruited 1,302 participants at the 14∼28th day after the second dose of inactivate SARS-CoV-2 vaccination and collected safety surveys. Among them we enrolled 1,266 who had completed all survey questions including demographic information, underlying medical conditions (disease type, duration, severity, and control status) as well as occurrence of adverse reaction events after each dose of vaccination. 297 individuals were excluded from this analysis due to receiving two different products of inactivated vaccine in each dose. The remaining 969 participants comprised the analyzed samples: 740 people with underlying medical condition and 229 people as healthy control. All of them formed the safety population. For the immunogenicity analysis, 969 participants had completed the blood sampling in the 14∼28th day after the second dose of vaccine. 178 participants failed to be on site and were excluded from the 90 days immunity assessment, and 40 participants with same reason were excluded from the 180 days analysis (Figure 1). Each disease group was further divided into 40∼59 years old and ≥60 years old subgroups with the baseline characteristics shown in the table 1.

**Table 1.** Demographic and clinical characteristics of the study participants.

|  | Healthy(N=229) |  | Comorbidities<br>(N=740) |  |
| --- | --- | --- | --- | --- |
|  | 40~59 years old<br>(N= 121) | ≥60 years old<br>(N=108) | 40~59 years old<br>(N=297) | ≥60 years old<br>(N=443) |
| Age, years | 51.50(5.55) | 66.89(5.04) | 53.63(5.34) | 70.28(6.54) |
| Height, m | 1.62(6.98) | 1.61(9.31) | 1.61(7.96) | 1.60(8.53) |
| Weight, kg | 60.20(9.96) | 61.16(11.75) | 64.90(12.62) | 60.95(10.31) |
| BMI, kg/m<sup>2</sup> | 22.83(3.10) | 23.47(3.42) | 24.83(3.97) | 23.71(3.55) |
| Sex, M/F | 55/66 | 42/66 | 152/141 | 187/256 |
| Han nationality | 121(100.00%) | 108(100.00%) | 285(95.96%) | 435(98.19%) |
| **Comorbidities** |  |  |  |  |
| Coronary artery disease (CAD) | - | - | 30(10.10%) | 88(19.86%) |
| Hypertension | - | - | 91(30.64%) | 141(31.83%) |
| Diabetes mellitus (DM) | - | - | 67(22.56%) | 110(24.83%) |
| Chronic respiratory disease (CRD) | - | - | 28(9.43%) | 66(14.90%) |
| Obesity | - | - | 16(5.39%) | 15(3.39%) |
| Cancer | - | - | 45(15.15%) | 43(9.71%) |

Data are mean (SD) or n (%) unless otherwise specified.

**Figure 1.**
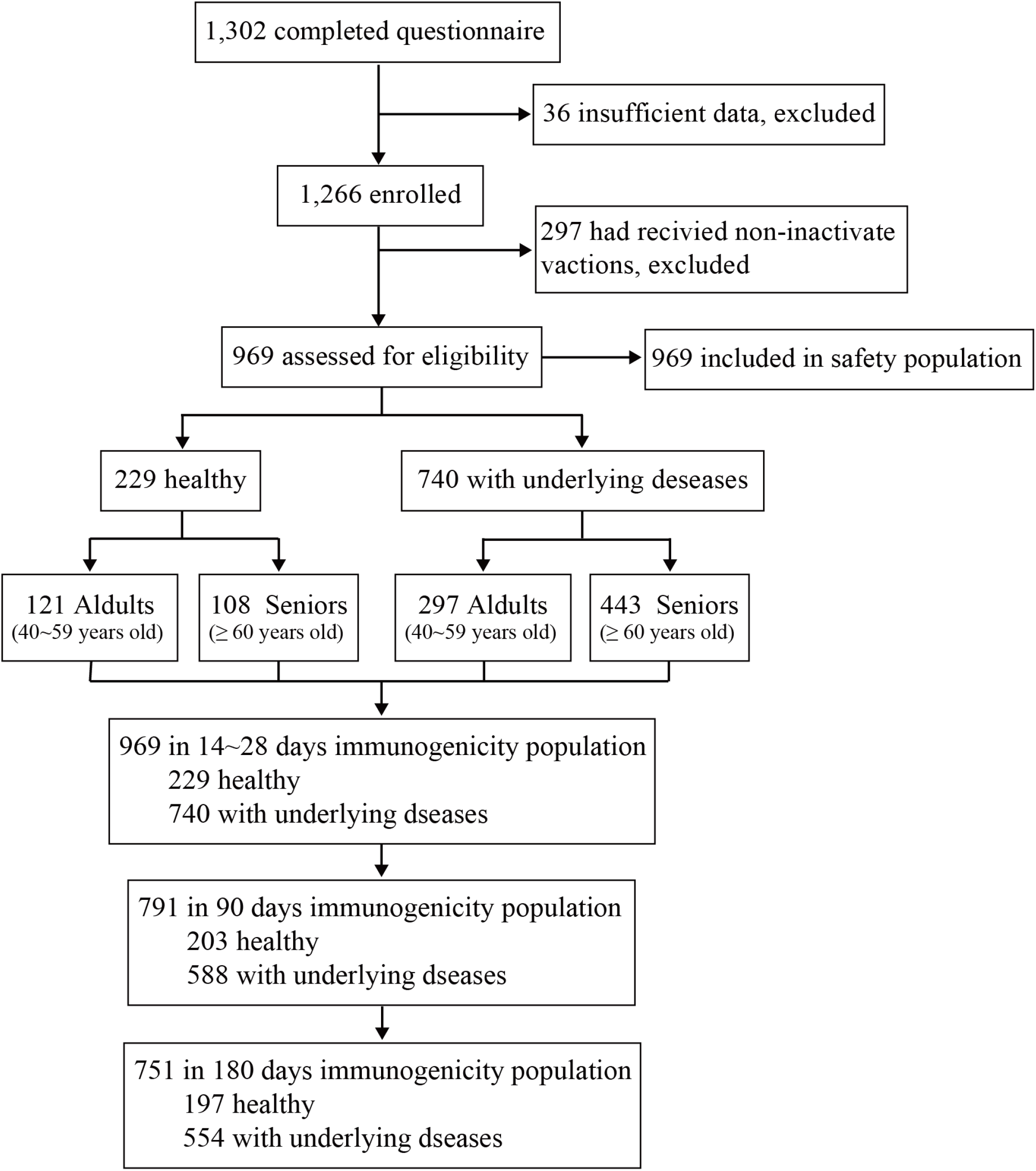
Study flow chart.

150 (20.27%) of 740 comorbidities participants had at least one adverse reaction, compared with 32 (13.97%) of 229 healthy participants (Appendix table 1). Most adverse events were mild (grade 1) in severity and participants recovered within 48 hours (Appendix table 2). The most frequently reported vaccine reactions in healthy and comorbidities group were injection-site pain (20 [8.73%] of 229 versus 94 [12.70%] of 740), fatigue (9 [3.93%] of 229 versus 48 [6.49%] of 740) and fever (0 [0.00%] of 229 versus 4 [0.54%] of 740). There was no significant difference seen between healthy and comorbidities cohort at overall level (Appendix table 1). Although 6 cases of grade 3 adverse events had happened in 4 individuals in comorbidities group, including acute allergy, skin & mucosa abnormalities and fever, which occurred 7 days after vaccination except for fever that occurred at the 1st day post vaccination. Thus, none was considered to be related to the vaccination except for the fever, which recovered at the 2nd day post vaccination (Appendix p.4-5).

**Table 2.** Incidence of adverse reactions after vaccination in healthy group and comorbidities group.

|  | Healthy<br>(N=229) |  | Comorbidities<br>(N=740) |  | Total<br>(N=969) |  | P Value<sup>[1]</sup> |  |
| --- | --- | --- | --- | --- | --- | --- | --- | --- |
|  | 40~59<br>years old<br>(N=121) | ≥60<br>years old<br>(N=108) | 40~59<br>years old<br>(N=297) | ≥60<br>years old<br>(N=443) | 40~59<br>years old<br>(N=418) | ≥60<br>years old<br>(N=551) | 40~59<br>years old | ≥60<br>years<br>old |
| Total adverse reactions | 20(0.17) | 12(0.11) | 81(0.27) | 69(0.16) | 101(0.24) | 81(0.15) | 0.0231 | 0.2895 |
| Local reactions | 17(0.14) | 5(0.05) | 61(0.21) | 41(0.09) | 78(0.19) | 46(0.08) | 0.1302 | 0.1722 |
| Pain | 15(0.12) | 5(0.05) | 59(0.20) | 35(0.08) | 74(0.18) | 40 (0.07) | 0.0893 | 0.3035 |
| Scleroma | 0(0.00) | 0(0.00) | 5(0.02) | 3(0.01) | 5(0.01) | 3(0.01) | 0.3275 | 1.0000 |
| Swelling | 3(0.02) | 0(0.00) | 5(0.02) | 5(0.01) | 8(0.02) | 5(0.01) | 0.6956 | 0.5886 |
| Redness | 0(0.00) | 0(0.00) | 2(0.01) | 1(0.00) | 2(0.00) | 1(0.00) | 1.0000 | 1.0000 |
| Rash | 0(0.00) | 0(0.00) | 2(0.01) | 1(0.00) | 2(0.00) | 1(0.00) | 1.0000 | 1.0000 |
| Itching | 0(0.00) | 0(0.00) | 3(0.01) | 6(0.01) | 3(0.01) | 6(0.01) | 0.5599 | 0.6031 |
| Systemic reactions | 6(0.05) | 7(0.06) | 40(0.13) | 33(0.07) | 46(0.11) | 40(0.07) | 0.0099 | 0.8383 |
| Acute allergy | 0(0.00) | 0(0.00) | 3(0.01) | 4(0.01) | 3(0.01) | 4(0.01) | 0.5599 | 1.0000 |
| Skin & mucosa<br>abnormalities | 0(0.00) | 0(0.00) | 1(0.00) | 4(0.01) | 1(0.00) | 4(0.01) | 1.0000 | 1.0000 |
| Diarrhoea | 1(0.01) | 0(0.00) | 2(0.01) | 1(0.00) | 3(0.01) | 1(0.00) | 1.0000 | 1.0000 |
| Anorexia | 0(0.00) | 1(0.01) | 4(0.01) | 1(0.00) | 4(0.01) | 2(0.00) | 0.3284 | 0.3539 |
| Vomiting | 0(0.00) | 0(0.00) | 2(0.01) | 1(0.00) | 2(0.00) | 1(0.00) | 1.0000 | 1.0000 |
| Nausea | 0(0.00) | 0(0.00) | 5(0.02) | 1(0.00) | 5(0.01) | 1(0.00) | 0.3275 | 1.0000 |
| Muscle pain | 1(0.01) | 0(0.00) | 12(0.04) | 6(0.01) | 13(0.03) | 6(0.01) | 0.1203 | 0.6031 |
| Headache | 0(0.00) | 1(0.01) | 7(0.02) | 5(0.01) | 7(0.02) | 6(0.01) | 0.2005 | 1.0000 |
| Cough | 1(0.01) | 0(0.00) | 4(0.01) | 2(0.00) | 5(0.01) | 2(0.00) | 1.0000 | 1.0000 |
| Fatigue | 3(0.02) | 6(0.06) | 26(0.09) | 22(0.05) | 29(0.07) | 28(0.05) | 0.0199 | 0.8075 |
| Fever | 0(0.00) | 0(0.00) | 2(0.01) | 2(0.00) | 2(0.00) | 2(0.00) | 1.0000 | 1.0000 |

<sup>[1]</sup> The p-value was calculated by Fisher's exact probability method.

When inspecting the adverse reactions after the first dose and the second dose of vaccination, respectively, the incidence of adverse events were 16 (6.99%) of 229 in the health group versus 97 (13.11%) of 740 in the comorbidities group after the first dose, 19 (8.30%) of 229 versus 99 (13.37%) of 740 after the second dose (Appendix table 1), with significant difference between health and comorbidities group in both doses (P=0.0129, first dose; P=0.0487, second dose). We then stratified participants by age group: adults (40-59 years old) and seniors (≥60 years old), to explore if seniors exhibit different response to inactivated SARS-CoV-2 vaccine. In the adults subgroup, 81 (27.27%) of 297 participants in the comorbidities group versus 20 (16.53%) of 121 participants in the health group, reported adverse events, with statistically significant difference between groups (P=0.0231). Moreover, systemic adverse reactions of the comorbidities group was significant higher than that of the health group (13.47% versus 4.96%, P=0.0099), while local adverse events showed no significant difference (Table 2). The incidence of adverse reactions was 54 (18.18%) of 297 in comorbidities people and 9 (7.44%) of 121 in healthy people after the first dose, and 55 (18.52%) of 297 in comorbidities people and 12 (9.92%) of 121 (9.92%) in healthy people after the second dose of vaccine. Both doses showed significant differences between diseases and healthy control (P=0.0062 and P=0.0387). In the senior subgroup, comorbidities and healthy control did not show significant difference in overall (15.58% versus 11.11%, P=0.2895), first dose (9.71% versus 6.48%, P=0.3537) or second dose (9.93% versus 6.48%, P=0.3543) vaccination. Thus, the slightly higher incidence of adverse reactions in the adult comorbidities population was the main reason driving the difference in the overall incidence of adverse reactions between the comorbidities cohort and healthy control. More specifically, the major inter-group difference was contributed by the systemic adverse reactions, mainly fatigue (26 [8.75%] of 297 in comorbidities, 3 [2.48%] of 121 in health, P=0.0199).

Next, we compared the incidence of adverse events between comorbidities and healthy control stratified by disease types (Appendix table 3). The overall incidence of adverse events was 46 (19.83%) of 232 in hypertension group, 24 (20.34%) of 118 in CAD group, 34 (19.21%) of 177 in DM group, 22 (23.40%) of 94 in CRD group, 19 (21.59%) of 88 in cancer group, 5 (6.13%) of 31 in obesity group versus 32 (13.97%) of 229 in health group. The most frequently reported adverse reactions in six disease groups were the same as those in the health group, mainly injection-site pain and fatigue (Figure 2). All adverse reactions showed no significant difference between six disease groups and health group. When focusing on seniors, CRD group showed significantly higher incidence of injection-site pain (9 [14.06%] of 64 vs. 5 [4.63%] of 108 in the health group, P=0.0404) than that of the healthy control. Regarding adults, fatigue in the hypertension group (11 [10.89%] of 101 versus 3 [2.48%] of 121 in health group, p=0.0124), DM group (7 [9.86%] of 71, P=0.0403) and cancer group (6 [12.77%] of 47, P=0.0152), and headache in the CRD group (2 [6.45%] of 31 versus 0 [0.00%] of 121 in health group, P=0.0405) showed significant differences compared to the healthy control.

**Table 3.**
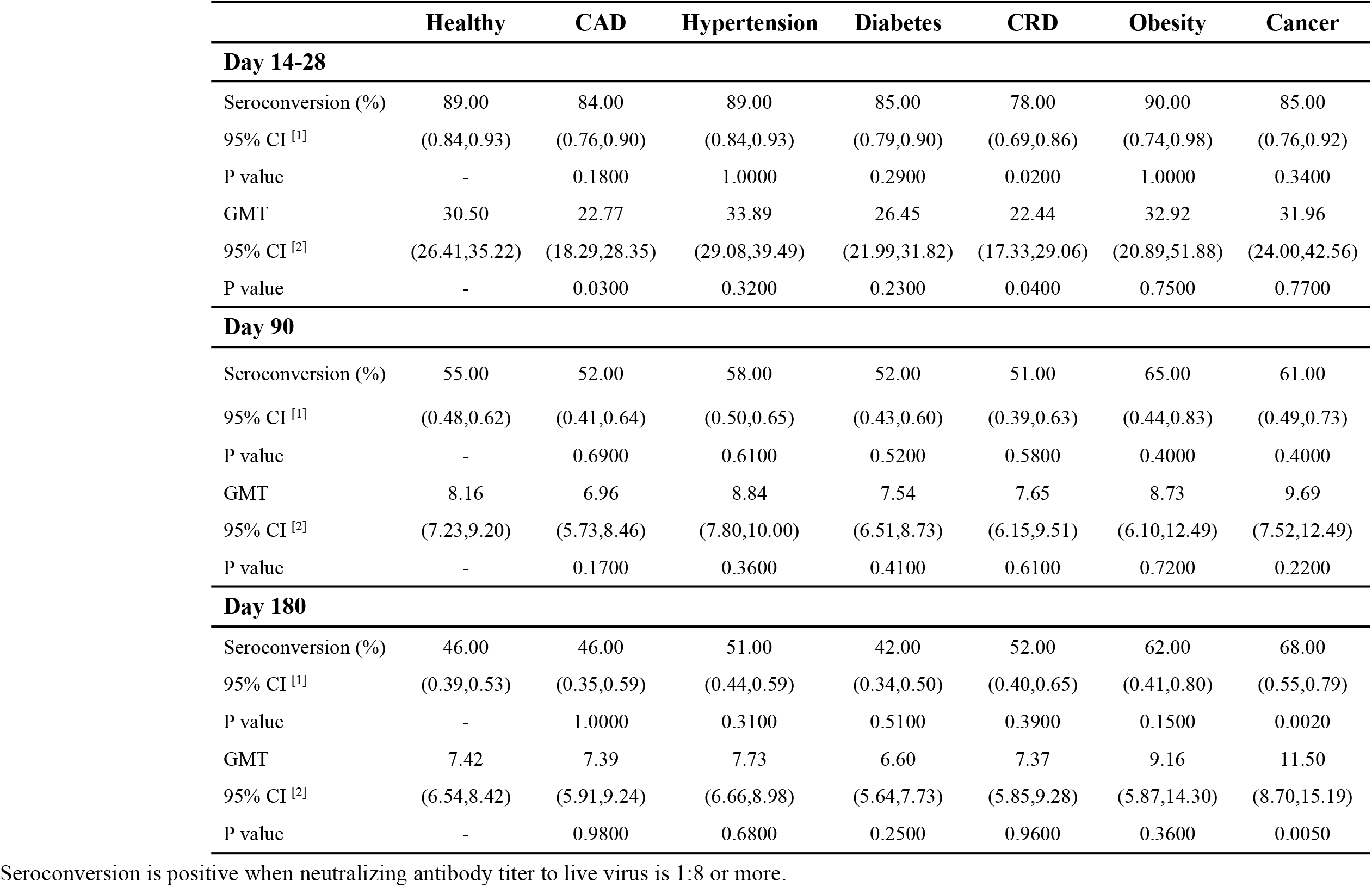
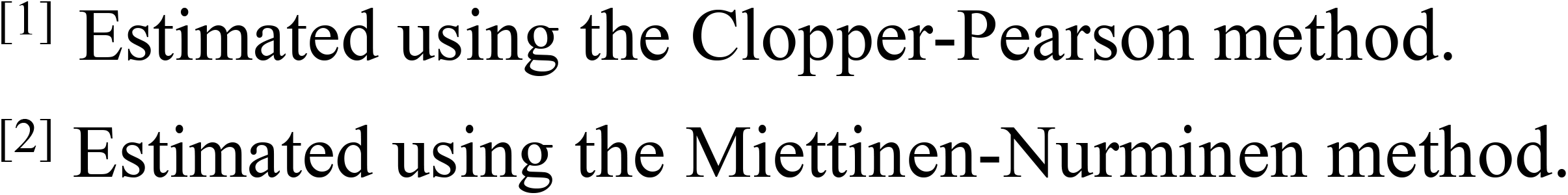
Immunogenicity among participants with underling disease and healthy 14-28 days, 3 months, and 6 months after full two-dose vaccination.

**Figure 2.**
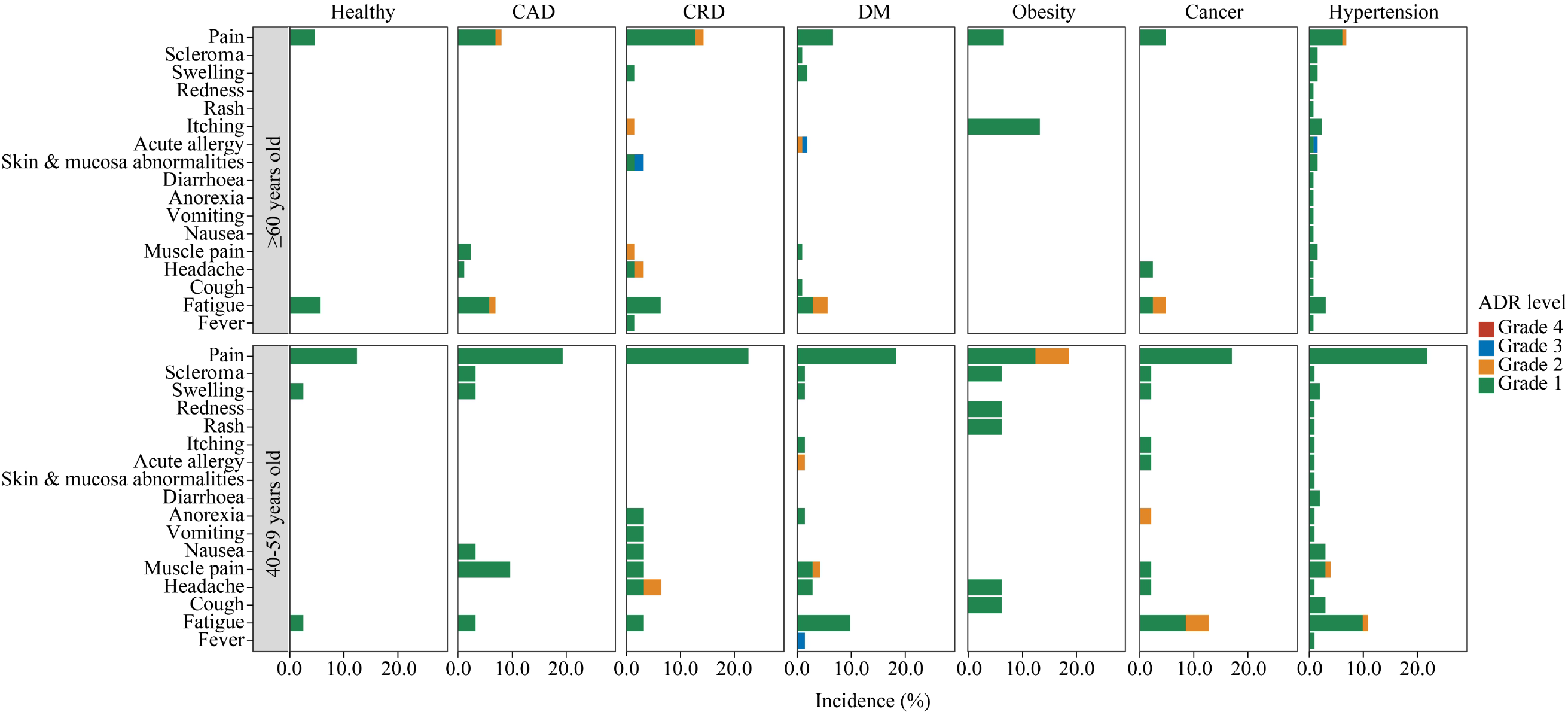
**Incidence of adverse reactions reported within 14 days post the first dose and the second dose of the vaccination in the safety population**.

The immunogenicity of people with underlying diseases and healthy control was evaluated at 14-28 days, 3 months, and 6 months after the full two-dose vaccination (Table 3). By day 14-28, the seroconversion rates of neutralizing antibodies were 99 (84%) of 118 in CAD group, 206 (89%) of 232 in hypertension group, 150 (85%) of 177 in DM group, 73 (78%) of 94 in CRD group, 28 (90%) of 31 in obesity group, 75 (85%) of 88 in cancer group versus 204 (89%) of 229 in health group; The neutralizing antibody GMTs were 22.7 (95% CI 18.29-28.35), 33.89 (95% CI 29.08-39.49), 26.45 (95% CI 21.99-31.82), 22.44 (95% CI 17.33-29.06), 32.92 (95% CI 20.89-51.88), 31.96 (95% CI 24.00-42.56) versus 30.50 (95% CI 26.41-35.22), respectively. Most diseases showed no difference in seroconversion rates and GMTs from healthy control while CAD (P=0.03) and CRD group (P=0.04) exhibited statistically significant decrease of humoral immune response. By day 90, both seroconversion rates and GMTs were significantly reduced in all people, with no difference between disease and health groups. By day 180, the seroconversion rates and GMTs continued to drop, but declined at a smaller scale. Interestingly, cancer patients showed a significant elevation in seroconversion rate (68% versus 46%, P=0.002) and GMT (11.50 versus 7.42, P=0.005) compared to healthy control (Table 3).

In the adult subgroups (40-59 years old), GMTs of neutralizing antibodies showed some elevations in cancer (53.71 versus 28.71, P=1.62 × 10^−3^) and hypertension (49.02 versus 28.71, P=8.9 × 10^−4^) patients compared to healthy control (Appendix table 4). Interestingly, in the senior subgroups (≥60 years old), cancer patients (18.56 versus 32.44, P=0.0200) showed the opposite trend with a significantly reduced GMT level. Similarly, senior CAD (20.05, P=0.0052) and CRD (19.85, P=0.0144) patients also showed the same trend of immunogenicity reduction (Appendix table 5). Notably, these inter-group variations were no longer significant at 3- and 6-months post vaccination.

Comparing across age groups, we observed distinct patterns for disease and health groups. In healthy participants, seroconversion and GMT of neutralizing antibodies were slightly higher in seniors (≥ 60 years old) than their younger counterpart (40-59 years old) on day 14-28 and 90 after the second dose. Conversely, senior people with underlying diseases had a lower neutralizing antibody level compared to their younger counterpart by day 14-28 post vaccination. These differences between age groups in seroconversion rate and GMT of neutralizing antibodies diminished by the time of 3 and 6 months after the second dose (Figure 3A). The reverse cumulative distribution plot showed that, at each time point, the overall distribution of neutralizing antibody titers are generally close across diseases in the senior and adult subgroups (Figure 3B). This suggests that the immunogenicity of CoronaVac inactivated vaccine in people with common chronic disease are generally as effective as that in healthy people.

**Figure 3.**
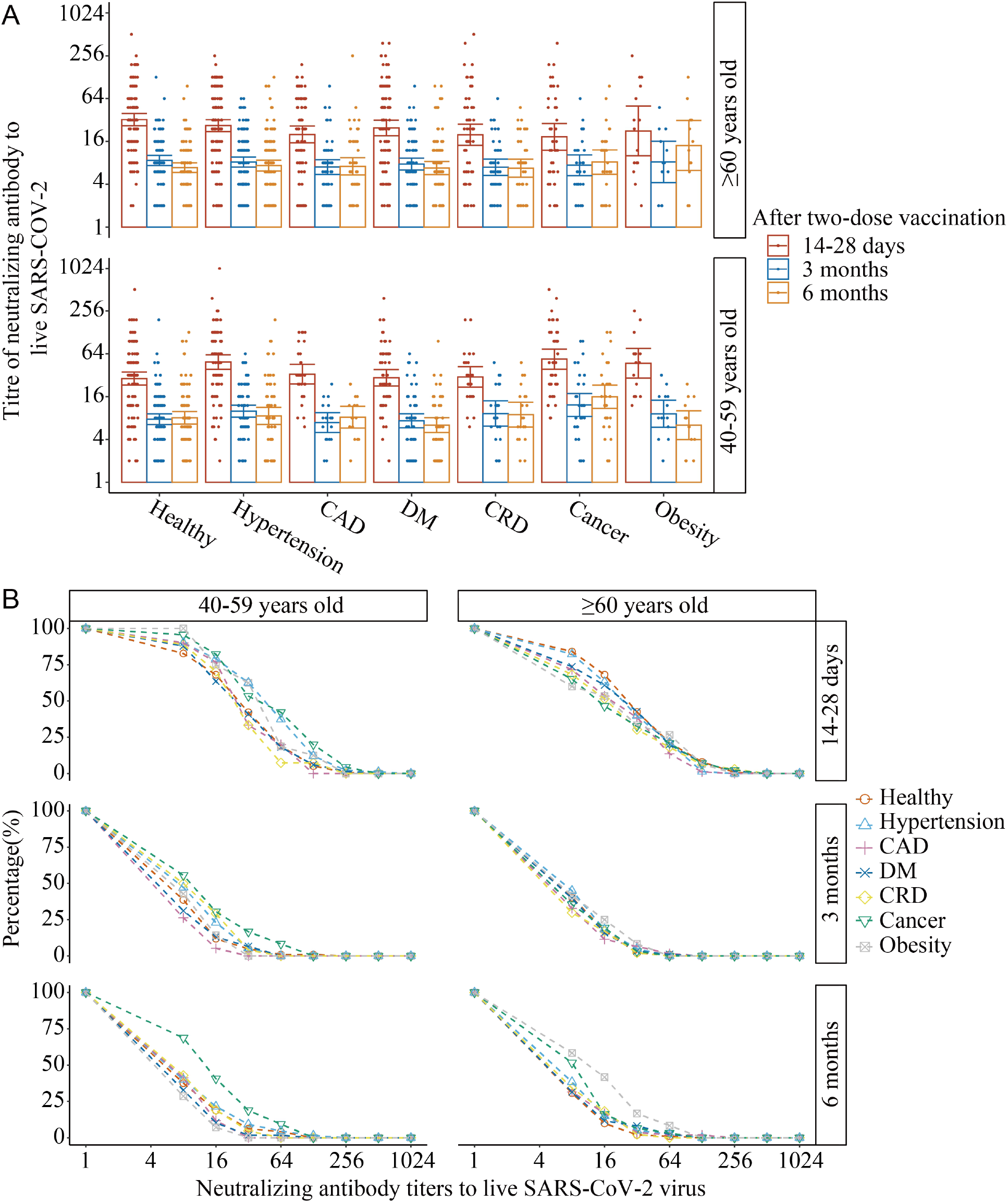
Neutralizing antibodies to live SARS-CoV-2 virus (wild type) induced 14-28 days, 90 days, and 180 days after two-dose CoronaVac. Antibody titers (A) and the inverse cumulative distribution (B) of neutralizing antibodies in different chronic diseases subgroups and healthy control, across age groups

## Discussion

People living with underlying medical condition, especially seniors, are at high risk for severe COVID-19-related complications and mortality. Considering the risk vs. benefit, vaccination against SARS-CoV-2 should be prioritized for this special population. Guidelines from the USA, the UK, the Korean and the WHO recommend COVID-19 vaccination for patients with comorbidities. However, safety data for the application of inactivated vaccine, the most widely administered vaccine type in China and some countries, in this vulnerable population has been missing. The lack of data on this special population partially contributed to the vaccine hesitancy and hence low coverage of vaccination among people aged 60 or older in China.

We present the first large cohort study addressing the safety and immunogenicity of CoronaVac inactivated vaccine among population living with underlying diseases compared with healthy control. For the safety, there was no significant difference in the incidence of adverse reactions between the underlying disease population and the healthy population in most of the diseases and age subgroups. For diseases at certain age group showing elevated incidence of adverse events, the differences were mainly contributed by injection-site pain and fatigue. In addition, these adverse events were mostly mild (grade 1), which could recover within 1-2 days without intervention.

For the immunogenicity, there were no significant differences in most of the disease and age subgroups compared with the healthy control. Although a few age and diseases subgroups had statistically significant lower immunogenicity than control, the differences were less than 40% and diminished over time. Thus, the immunogenicity of CoronaVac in people with chronic diseases, particularly in people over 60 years of age, was comparable with healthy counterparts. More importantly, the general trend of slightly lower antibody response in seniors emphasized the importance of this population to receive booster doses upon completion of primary immunization.

For the widely used COVID-19 vaccines, some had recruited participants with underlying medical conditions in the phase 3 clinical trials. For example, the Pfizer COVID-19 mRNA vaccine (BNT162b2) included 20.3% of the sample population reporting one or more underlying diseases. The most common comorbidities were hypertension (24.5%), diabetes mellitus (DM) (7.8%), and chronic lung disease (7.8%). Patients with comorbidities showed similar preventive results and adverse events with the total sample population^26,27^. In the phase 3 clinical trials (P301) for the Moderna COVID-19 vaccine (mRNA-1273), 22.2% of the sample population had high-risk underlying medical conditions. The most common comorbidities were DM (9.4%), severe obesity with body mass index (BMI) ≥40 kg/m^2^ (6.5%), severe heart disease (4.9%), and CRD (4.8%). The incidence of adverse events was similar in the group with comorbidities and the total sample population with comparable preventive effects^27,28^. Our analysis for CoronaVac, the most widely used inactivated SARS-COV-2 vaccine, showed consistent result with those analyzed in mRNA vaccine trials. Our result for the first time, provided a comprehensive picture of the immunogenicity and safety of COVID-19 inactivated vaccines in people (especially seniors) with underlying medical conditions.

Our study has a few limitations. First, the safety data of this retrospective study was collected 14-28 days after vaccination instead of daily report. The accuracy of adverse event reports could thus be somewhat affected. Second, we observed a significant reduction of neutralizing antibody titers (humoral immunity) 3 months after the second dose, but the duration of cellular immunity at this time point remains unclear. Our ongoing efforts aim to evaluate the cellular immunity against SARS-CoV-2 at the 3rd and 6th months post vaccination. Third, limited by the sample size, we only considered the rough classification of the chronic diseases for highly heterogeneous diseases. For example, some subtypes of cancer may have great impact on the immune system, resulting in an abnormal response to the vaccination. However, the assessment for safety and immunogenicity of CoronaVac on HIV positive or autoimmune rheumatic diseases (ARD) patients showed that the immunogenicity of immunosuppressed patients was significantly decreased, but remained within an acceptable range; and they all showed a good safety profile with the CoronaVac vaccines^29,30^. Therefore, we believe that CoronaVac will be safe in immune-related cancers but requires further efficacy and safety evaluation to provide precise vaccination guidance. In conclusion, CoronaVac vaccination showed similar efficacy and safety in individuals with and without common underlying medical conditions. Considering that individuals with comorbidities have a significantly higher risk of progression to severe conditions or death when infected with SARS-CoV-2, the benefits of the COVID-19 vaccination outweigh the risks. Thus, we strongly recommend people with common chronic diseases get vaccinated as soon as possible given their disease status were stable.

## Supporting information

Appendix table 1-5 & Serious adverse events reported during the study

## Data Availability

De-identified data are freely available from the corresponding author upon request.

## Contributors

Z.Z., C.L., Z.F. designed and supervised this study. Z.Z. C.L. wrote the manuscript. A.L., H.B. performed the experiments and analysis. All other authors participated in patient screening, sample and data collection.

## Data sharing

De-identified data are freely available from the corresponding author upon request.

## Acknowledgments

We thank the Yunnan Provincial Science and Technology Department (202102AA100051 and 202003AC100010, China) and Sinovac Biotech Ltd (PRO-nCOV-4004) for research funding. We thank researchers and clinicians from Haikou, Wenchang and Qionghai CDC for generous support on volunteer recruitment.

