## Appendix table 1-5 & Serious adverse events reported during the study for "Immunogenicity and safety of the CoronaVac inactivated SARS-CoV-2 vaccine in people with underlying medical conditions: a retrospective study"

Table 1. Incidence of adverse events after vaccination in health group and comorbidities group.

| **Adverse reactions** | **Health** | | **Comorbidities** | | **Total** | | **P-value ^[1]^** |
| --- | --- | --- | --- | --- | --- | --- | --- |
|  | **(N=229)** | | **(N=740)** | | **(N=969)** | | **Health vs.**  **Comorbidities** |
|  | **Cases** | **n ^[2]^ (%)** | **Cases** | **n ^[2]^ (%)** | **Cases** | **n ^[2]^ (%)** |  |
| Any adverse reactions | 39 | 32 (13.97) | 322 | 150 (20.27) | 361 | 182 (18.78) | 0.0334 |
| Systemic reactions | 15 | 13 (5.68) | 161 | 73 (9.86) | 176 | 86 (8.88) | 0.0618 |
| Local reactions | 24 | 22 (9.61) | 161 | 102 (13.78) | 185 | 124 (12.80) | 0.1129 |
| First dose | 18 | 16 (6.99) | 158 | 97 (13.11) | 176 | 113 (11.66) | 0.0129 |
| Second dose | 21 | 19 (8.30) | 164 | 99 (13.38) | 185 | 118 (12.18) | 0.0487 |

[1] The P-value was calculated by Fisher's exact test.
[2] Number of participants reporting at least 1 occurrence of the specified event category.

Table 2. Incidence of adverse events after any vaccination dose for all participants stratified by severity of adverse events.

| **Adverse reactions** | **Grade 1** | **Grade 2** | **Grade 3** | **Total** |
| --- | --- | --- | --- | --- |
| Any adverse reactions | 206 (16.3%) | 19 (1.4%) | 4 (0.3%) | 229 (18.1%) |
| Local reactions | 158 (12.5%) | 5 (0.4%) | 0 (0%) | 163 (12.9%) |
| Pain | 147 (11.6%) | 4 (0.3%) | 0 (0%) | 151 (11.9%) |
| Scleroma | 9 (0.7%) | 0 (0%) | 0 (0%) | 9 (0.7%) |
| Swelling | 14 (1.1%) | 0 (0%) | 0 (0%) | 14 (1.1%) |
| Redness | 3 (0.2%) | 0 (0%) | 0 (0%) | 3 (0.2%) |
| Rash | 3 (0.2%) | 0 (0%) | 0 (0%) | 3 (0.2%) |
| Itching | 8 (0.6%) | 1 (0.1%) | 0 (0%) | 9 (0.7%) |
| Systemic reactions | 77 (6%) | 15 (1.2%) | 4 (0.3%) | 96 (7.6%) |
| Acute allergy | 3 (0.2%) | 2 (0.2%) | 2 (0.2%) | 7 (0.6%) |
| Skin & mucosa abnormalities | 4 (0.3%) | 0 (0%) | 1 (0.1%) | 5 (0.4%) |
| Diarrhoea | 4 (0.3%) | 0 (0%) | 0 (0%) | 4 (0.3%) |
| Anorexia | 5 (0.4%) | 2 (0.2%) | 0 (0%) | 7 (0.6%) |
| Vomiting | 3 (0.2%) | 0 (0%) | 0 (0%) | 3 (0.2%) |
| Nausea | 6 (0.5%) | 1 (0.1%) | 0 (0%) | 7 (0.6%) |
| Muscle pain | 16 (1.3%) | 3 (0.2%) | 0 (0%) | 19 (1.4%) |
| Headache | 11 (0.9%) | 2 (0.2%) | 0 (0%) | 13 (1.0%) |
| Cough | 8 (0.6%) | 0 (0%) | 0 (0%) | 8 (0.6%) |
| Fatigue | 57 (4.5%) | 8 (0.6%) | 0 (0%) | 65 (5.1%) |
| Fever | 3 (0.2%) | 0 (0%) | 1 (0.1%) | 4 (0.%) |

Table 3. Incidence of adverse events after vaccination in health group and different disease groups in the adults (40-59 years old) and seniors (≥60 years old) cohort.

|  | **Healthy (N=229)** | | **Hypertension (N=232)** | | **Total (N=461)** | | **P-value** | |
| --- | --- | --- | --- | --- | --- | --- | --- | --- |
|  | **40~59 years old (N=121)** | **≥60 years old (N=108)** | **40~59 years old(N=101)** | **≥60 years old (N=131)** | **40~59 years old(N=222)** | **≥60 years old(N=239)** | **40~59 years old** | **≥60 years old** |
| Total adverse reactions | 20 (17%) | 12 (11%) | 29 (29%) | 17 (13%) | 49 (22%) | 29 (12%) | 0.0349 | 0.6954 |
| Local reactions | 17 (14%) | 5 (5%) | 22 (22%) | 11 (8%) | 39 (18%) | 16 (7%) | 0.1574 | 0.3040 |
| Systemic reactions | 6 (5%) | 7 (6%) | 15 (15%) | 7 (5%) | 21 (9%) | 14 (6%) | 0.0195 | 0.7856 |
|  | **Healthy (N=229)** | | **CAD (N=118)** | | **Total (N=347)** | | **P-value** | |
|  | **40~59 years old (N=121)** | **≥60 years old (N=108)** | **40~59 years old (N=31)** | **≥60 years old (N=87)** | **40~59 years old (N=152)** | **≥60 years old (N=195)** | **40~59 years old** | **≥60 years old** |
| Total adverse reactions | 20 (17%) | 12 (11%) | 8 (26%) | 16 (18%) | 28 (18%) | 28 (14%) | 0.2977 | 0.1574 |
| Local reactions | 17 (14%) | 5 (5%) | 6 (19%) | 7 (8%) | 23 (15%) | 12 (6%) | 0.5737 | 0.3778 |
| Systemic reactions | 6 (5%) | 7 (6%) | 4 (13%) | 9 (10%) | 10 (7%) | 16 (8%) | 0.1207 | 0.4326 |
|  | **Healthy (N=229)** | | **DM (N=177)** | | **Total (N=406)** | | **P-value** | |
|  | **40~59 years old (N=121)** | **≥60 years old (N=108)** | **40~59 years old (N=71)** | **≥60 years old (N=106)** | **40~59 years old (N=192)** | **≥60 years old (N=214)** | **40~59 years old** | **≥60 years old** |
| Total adverse reactions | 20 (17%) | 12 (11%) | 19 (27%) | 15 (14%) | 39 (20%) | 27 (13%) | 0.0974 | 0.5421 |
| Local reactions | 17 (14%) | 5 (5%) | 13 (18%) | 9 (8%) | 30 (16%) | 14 (7%) | 0.5372 | 0.2819 |
| Systemic reactions | 6 (5%) | 7 (6%) | 9 (13%) | 8 (8%) | 15 (8%) | 15 (7%) | 0.0912 | 0.7950 |
|  | **Healthy (N=229)** | | **CRD (N=94)** | | **Total (N=323)** | | **P-value** | |
|  | **40~59 years old (N=121)** | **≥60 years old (N=108)** | **40~59 years old (N=31)** | **≥60 years old (N=63)** | **40~59 years old (N=222)** | **≥60 years old (N=239)** | **40~59 years old** | **≥60 years old** |
| Total adverse reactions | 20 (17%) | 12 (11%) | 8 (26%) | 14 (22%) | 49 (22%) | 29 (12%) | 0.2977 | 0.0758 |
| Local reactions | 17 (14%) | 5 (5%) | 7 (23%) | 10 (16%) | 39 (18%) | 16 (7%) | 0.2721 | 0.0219 |
| Systemic reactions | 6 (5%) | 7 (6%) | 3 (10%) | 6 (10%) | 21 (9%) | 14 (6%) | 0.3893 | 0.5533 |
|  | **Healthy (N=229)** | | **Cancer (N=88)** | | **Total (N=317)** | | **P-value** | |
|  | **40~59 years old (N=121)** | **≥60 years old (N=108)** | **40~59 years old (N=47)** | **≥60 years old (N=41)** | **40~59 years old (N=168)** | **≥60 years old (N=149)** | **40~59 years old** | **≥60 years old** |
| Total adverse reactions | 20 (17%) | 12 (11%) | 14 (30%) | 5 (12%) | 34 (20%) | 17 (11%) | 0.0853 | 1.0000 |
| Local reactions | 17 (14%) | 5 (5%) | 10 (21%) | 2 (5%) | 27 (16%) | 7 (5%) | 0.2520 | 1.0000 |
| Systemic reactions | 6 (5%) | 7 (6%) | 8 (17%) | 3 (7%) | 14 (8%) | 10 (7%) | 0.0244 | 1.0000 |
|  | **Healthy (N=229)** | | **Obesity (N=31)** | | **Total (N=260)** | | **P-value** | |
|  | **40~59 years old (N=121)** | **≥60 years old (N=108)** | **40~59 years old (N=16)** | **≥60 years old (N=15)** | **40~59 years old (N=137)** | **≥60 years old (N=123)** | **40~59 years old** | **≥60 years old** |
| Total adverse reactions | 20 (17%) | 12 (11%) | 3 (19%) | 2 (13%) | 23 (17%) | 14 (11%) | 0.7328 | 0.6799 |
| Local reactions | 17 (14%) | 5 (5%) | 3 (19%) | 2 (13%) | 20 (15%) | 7 (6%) | 0.7048 | 0.2033 |
| Systemic reactions | 6 (5%) | 7 (6%) | 1 (6%) | 0 (0%) | 7 (5%) | 7 (6%) | 0.5895 | 0.5959 |

Table 4. Seroconversion rate and geometric mean titer (GMT) in the health group and different disease groups in the adults (40-59 years old) cohort.

|  |  | **Healthy** | **CAD** | **Hypertension** | **Diabetes** | **CRD** | **Obesity** | **Cancer** |
| --- | --- | --- | --- | --- | --- | --- | --- | --- |
| **Day 14-28** | Seroconversion (%) | 86.00 | 97.00 | 93.00 | 92.00 | 96.00 | 100.00 | 100.00 |
|  | 95% CI | (0.79,0.92) | (0.83,1.00) | (0.86,0.98) | (0.83,0.97) | (0.81,1.00) | (0.83,1.00) | (0.94,1.00) |
|  | P-value | - | 0.2000 | 0.1100 | 0.2400 | 0.2000 | 0.2200 | 0.0064 |
|  | GMT | 28.71 | 33.08 | 49.02 | 29.44 | 30.28 | 47.10 | 53.71 |
|  | 95% CI | (23.31,35.37) | (24.11,45.39) | (38.73,62.03) | (22.55,38.43) | (21.76,42.14) | (29.08,76.29) | (38.84,74.28) |
|  | P-value | - | 0.4500 | 0.0009 | 0.8800 | 0.7800 | 0.0600 | 0.0016 |
| **Day 90** | Seroconversion (%) | 53.00 | 58.00 | 65.00 | 51.00 | 62.00 | 71.00 | 72.00 |
|  | 95% CI | (0.43,0.63) | (0.33,0.80) | (0.53,0.76) | (0.38,0.64) | (0.41,0.81) | (0.42,0.92) | (0.55,0.86) |
|  | P-value | - | 0.8000 | 0.1600 | 0.7500 | 0.5000 | 0.2600 | 0.0800 |
|  | GMT | 7.70 | 6.89 | 9.97 | 7.31 | 9.25 | 9.17 | 12.21 |
|  | 95% CI | (6.45,9.18) | (5.00,9.51) | (8.23,12.07) | (5.81,9.21) | (6.13,13.95) | (5.92,14.19) | (8.42,17.69) |
|  | P-value | - | 0.5400 | 0.0500 | 0.7300 | 0.4100 | 0.4400 | 0.0300 |
| **Day 180** | Seroconversion (%) | 49.00 | 59.00 | 57.00 | 41.00 | 52.00 | 50.00 | 81.00 |
|  | 95% CI | (0.39,0.59) | (0.33,0.82) | (0.44,0.69) | (0.29,0.55) | (0.30,0.74) | (0.23,0.77) | (0.64,0.93) |
|  | P-value | - | 0.6000 | 0.3400 | 0.4100 | 0.8100 | 1.0000 | 0.0017 |
|  | GMT | 8.05 | 8.22 | 8.55 | 6.35 | 8.91 | 6.35 | 15.92 |
|  | 95% CI | 6.57,9.86 | (5.79,11.69) | (6.46,11.32) | (5.00,8.07) | (5.97,13.30) | (3.98,10.13) | (10.89,23.27) |
|  | P-value | - | 0.9100 | 0.7300 | 0.1400 | 0.6400 | 0.3300 | 0.0023 |

Table 5. Seroconversion rate and geometric mean titer (GMT) in the health group and different disease groups in the seniors (≥60 years old) cohort.

|  |  | **Healthy** | **CAD** | **Hypertension** | **Diabetes** | **CRD** | **Obesity** | **Cancer** |
| --- | --- | --- | --- | --- | --- | --- | --- | --- |
| **Day 14-28** | Seroconversion (%) | 92.00 | 80.00 | 86.00 | 81.00 | 71.00 | 80.00 | 70.00 |
|  | 95% CI | (0.85,0.96) | (0.70,0.87) | (0.79,0.91) | (0.72,0.88) | (0.59,0.82) | (0.52,0.96) | (0.54,0.83) |
|  | P-value | - | 0.0123 | 0.1600 | 0.0200 | 0.0004 | 0.1500 | 0.0012 |
|  | GMT | 32.44 | 20.05 | 26.70 | 24.81 | 19.85 | 22.46 | 18.56 |
|  | 95% CI | (26.53,39.67) | (15.29,26.30) | (22.02,32.39) | (19.30,31.88) | (14.15,27.86) | (10.04,50.25) | (12.04,28.61) |
|  | P-value | - | 0.0052 | 0.1700 | 0.1000 | 0.0144 | 0.3600 | 0.0200 |
| **Day 90** | Seroconversion (%) | 57.00 | 51.00 | 53.00 | 52.00 | 45.00 | 58.00 | 48.00 |
|  | 95% CI | (0.47,0.67) | (0.38,0.64) | (0.44,0.62) | (0.41,0.63) | (0.30,0.60) | (0.28,0.85) | (0.30,0.67) |
|  | P-value | - | 0.5200 | 0.6800 | 0.5600 | 0.2200 | 1.0000 | 0.4200 |
|  | GMT | 8.64 | 6.98 | 8.20 | 7.69 | 6.94 | 8.24 | 7.42 |
|  | 95% CI | (7.32,10.20) | (5.49,8.88) | (6.97,9.65) | (6.34,9.34) | (5.34,9.01) | (4.22,16.08) | (5.31,10.36) |
|  | P-value | - | 0.1500 | 0.6600 | 0.3700 | 0.1600 | 0.8800 | 0.4100 |
| **Day 180** | Seroconversion (%) | 43.00 | 43.00 | 48.00 | 42.00 | 52.00 | 75.00 | 55.00 |
|  | 95% CI | (0.33,0.53) | (0.29,0.57) | (0.39,0.58) | (0.31,0.53) | (0.37,0.68) | (0.43,0.95) | (0.36,0.73) |
|  | P-value | - | 1.0000 | 0.4200 | 1.0000 | 0.3600 | 0.0600 | 0.3000 |
|  | GMT | 6.87 | 7.14 | 7.32 | 6.77 | 6.73 | 14.05 | 8.22 |
|  | 95% CI | (5.88,8.02) | (5.41,9.43) | (6.15,8.72) | (5.47,8.37) | (5.04,8.99) | (6.26,31.55) | (5.54,12.17) |
|  | P-value | - | 0.8000 | 0.5900 | 0.9200 | 0.9000 | 0.0800 | 0.3900 |

### Serious adverse events reported during the study:

4 participants in this study had 6 cases of severe (grade 3) adverse events:

Individual 1, 60-65 years old, received the first dose of the vaccine on July, 2021, and the second dose on August, 2021. Grade 3 Skin & mucosa abnormalities (mouth ulcers) was found post 14 days of both doses of vaccination and recovered within one week. In addition, there were grade 2 headache after the first dose; and grade 2 injection-site pain, grade 1 swelling, grade 2 headache, grade 1 fatigue and grade 1 fever after the second dose. He (She) was enrolled in CRD group, with hypertension, cancer, and hyperthyroidism. His (Her) health condition is stable within 6 months before the first dose of vaccination. Since mouth ulcers are not highly associated with common adverse effects of vaccination, it is considered not associated with vaccination.

Individual 2, 80-85 years old, received the first dose of the vaccine on August, 2021, and the second dose of the vaccine on September, 2021. Only a grade 3 acute allergic reaction occurred after the second dose (October), which was manifested as systemic urticaria. He (She) was enrolled in hypertension group. His (Her) blood pressure is stable within 6 months before the first dose of vaccination. Follow-up visits were conducted every half a month, and as of February, 2022, he (she) still had symptoms of urticaria, which were relieved by taking drugs to treat urticaria. Considering that urticaria is an immune disease, it may be the cause of the immune system disorder in the body after vaccination, but because the onset time is more than 7 days from the vaccination, the possibility of seasonality and other factors cannot be ruled out.

Individual 3, 70-15 years old, received their first dose of the vaccine on September, 2021, and the second dose of the vaccine on September, 2021. Grade 3 acute allergic reaction occurred after both doses, specifically systemic urticaria. In addition, after the first dose of vaccination, there was grade 1 swelling at the injection-site; and after the second dose, there were grade 2 fatigue. He (She) was enrolled in DM group, with hypertension. His (Her) health condition is stable within 6 months before the first dose of vaccination, usually controlled by exercise and diet in addition to drugs. Considering that urticaria is an immune disease, it may be the cause of the immune system disorder in the body after vaccination, but because the onset time is more than 7 days from the vaccination, the possibility of seasonality and other factors cannot be ruled out.

Individual 4, 46-50 years old, received their first dose of the vaccine on June, 2021 and the second dose of the vaccine on July, 2021. Grade 3 fever occurred after the second dose of vaccination, with a maximum body temperature of 38.5℃, and recovered after taking the medicine. In addition, after the first dose of vaccination, there are grade 1 injection-site pain, grade 1 swelling, grade 1 anorexia, grade 1 muscle pain, grade 1 headache, and grade 1 fatigue; and after the second dose of vaccination, there are also grade 1 muscle pain, grade 1 headache and fatigue. He (She) was enrolled in DM group, with hypertension. The fever is associated with vaccination, but this is a common adverse vaccine reaction and was cured after drug therapy.
